# Redressing long-term antidepressant use (RELEASE): Pragmatic cluster randomised controlled trial in general practice

**DOI:** 10.64898/2026.08.19.26360323

**Authors:** Katharine A Wallis, Maria Donald, Mark Horowitz, Nicholas Zwar, Robert S Ware, Ian Scott, Christopher R Freeman, Maryanne Cleetus, Karen Thrift, Suzanne McDonald, Joanna Moncrieff

## Abstract

**BACKGROUND:** Safe and effective antidepressant deprescribing strategies are needed in general practice where most antidepressant prescribing occurs.

**METHODS:** We conducted a pragmatic, cluster-randomised controlled trial in general practice to test invitation to general practitioner (GP) review combined with resources to inform shared decision-making and guide hyperbolic tapering for stopping antidepressants compared to usual care. Adults taking antidepressants for longer than 12 months were recruited from 26 Australian GP practices between March 2023 and November 2024, irrespective of their intention to stop or depression or anxiety symptom scores. The primary outcome was cessation at 12 months. Secondary outcomes included cessation at 6 months, and ≥75% dose reduction and depression, anxiety and withdrawal symptom scores at 6 and 12 months.

**RESULTS:** Overall, 483 patients were randomised. Mean age was 50 years; 73% were women; mean duration of antidepressant use was 14.1 years. Cessation at 12 months was observed in 32 of 215 (14.9%) intervention and 16 of 187 (8.6%) usual care patients (odds ratio (OR) = 1.95 [95%CI, 1.00 to 3.81]; p=0.050). Cessation at 6 months was observed in 11.7% intervention vs 4.8% usual care (OR = 2.68; 95%CI, 1.18 to 6.05), and ≥75% dose reduction at 12 months in 19.6% intervention vs 9.9% usual care (OR = 2.28; 95%CI, 1.20 to 4.31). Symptom scores were similar between groups. No adverse events were attributable to the intervention.

**CONCLUSIONS:** In general practice, invitation to GP antidepressant review combined with information and guidance on hyperbolic tapering can support cessation or dose reduction without causing adverse effects or relapse. Absolute cessation rates were modest but still meaningful given the high prevalence of long-term antidepressant use.

**TRIAL REGISTRATION:** ANZCT registry identifier, ACTRN12622001379707p.

**WHAT IS ALREADY KNOWN ON THIS TOPIC:**

- Long-term (>12 months) antidepressant use is common in primary care, despite being potentially unnecessary and associated with increased risk of falls, sexual dysfunction, weight gain and diabetes.
- Lack of recognition and management of antidepressant withdrawal symptoms and fear of relapse are important drivers of long-term use.
- Safe and effective antidepressant deprescribing strategies are needed in primary care where most antidepressant prescribing occurs.

**WHAT THIS STUDY ADDS:**

- The RELEASE intervention comprised invitation to GP antidepressant review combined with patient-centred resources to inform shared decision-making and guide hyperbolic tapering.
- The intervention increased cessation or dose reduction, without increasing adverse effects including relapse, in a challenging population with mean duration of antidepressant therapy of 14.1 years and no intention to stop.

## Introduction

Safe, effective and scalable antidepressant deprescribing strategies are needed in primary care to address high and rising prevalence of long-term (>12 months) antidepressant use. There is a lack of high-quality evidence showing benefit with long-term use and some evidence showing harm including increased risk of diabetes, weight gain, falls and sexual dysfunction.^1–3^ Clinical guidelines generally recommend reviewing and considering deprescribing after six to twelve months.^4^ In the United Kingdom (UK), one in six people takes an antidepressant, of whom half take antidepressants for more than two years.^5, 6^ Comparable patterns are seen across other high-income countries, with use also higher among older people, women, and people living in socioeconomically disadvantaged areas.^7–11^

Most antidepressant prescribing occurs in primary care (80-92%),^10^ but effective deprescribing strategies in this clinical context are not known.^12–16^ There are multiple barriers to deprescribing in primary care,^17, 18^ despite evidence that even brief interventions can have some effect.^19^ There are additional drug-related barriers to deprescribing antidepressants including the belief that depression is a long-term condition caused by a chemical imbalance that antidepressants can correct, fear of relapse, and poor recognition and management of withdrawal symptoms.^20–23^ Withdrawal symptoms may arise following abrupt cessation or rapid dose reduction, and can be severe particularly among long-term users preventing cessation even in highly motivated patients.^24^ Animal studies suggest withdrawal symptoms may reflect neuroadaptation and rebound surge in neural excitability after cessation,^25^ mirroring patterns observed in other psychotropic withdrawal states. Hyperbolic tapering is recommended to minimize withdrawal symptoms.^26^ This approach involves progressively smaller dose reductions, to give steady decrease in serotonin transporter receptor occupancy, including through very low doses to eventual cessation.^27, 28^ Cohort studies indicate that this approach can support successful antidepressant cessation but controlled trials are lacking.^29^

We co-developed with people with lived experience of long-term antidepressant use and withdrawal symptoms patient-centred resources to support informed shared decision-making about whether to continue or stop taking antidepressants and guide hyperbolic tapering for stopping (www.releasetoolkit.com.au).^30, 31^ We conducted the Redressing long-term antidepressant use (RELEASE) trial in general practice to evaluate invitation to general practitioner (GP) review combined with the RELEASE resources for stopping long-term antidepressants compared to usual care.^32^ During the period this trial was conducted, hyperbolic tapering was a novel concept in Australian general practice, with no supportive resources and negligible implementation.^33^

## Methods

### Trial design and oversight

The study was a pragmatic, cluster randomised controlled trial (RCT) with 2-arms: intervention and usual care, clustered at the level of the general practice to avoid contamination between arms.^32^ Nested within the intervention arm, clusters were further randomised to RELEASE or RELEASE+ (a more intensive version of RELEASE). The primary objective was to assess the effectiveness of the RELEASE intervention (RELEASE and RELEASE+ arms combined) compared to usual care in supporting antidepressant cessation.^34^ The secondary objective was to assess any additional effectiveness of RELEASE+ compared to RELEASE alone.^34^ The protocol and statistical analysis plan have been published previously along with details of the intervention.^32, 34^

The study received approval from The University of Queensland Human Research Ethics Committee (2022/HE001667). All patients provided written informed consent. The trial was conducted in compliance with all International Council for Harmonisation Good Clinical Practice guidelines. A data and safety monitoring committee was established to monitor and review adverse events. This report followed the 2025 Consolidated Standards of Reporting Trials (CONSORT) reporting guidelines.

### Setting and participants

General practices in southeast Queensland, Australia were recruited between November 2022 and September 2024. Practices were identified from a list of practices providing clinical placements for medical students and invited by email to express their interest in participating, followed up via phone call. Local Primary Health Networks and GP education providers also promoted the trial via their newsletter. Practices were excluded if they did not have compatible practice management software. The trial practice liaison officer visited interested practices and obtained signed agreement to participate from practice principals. In Australia, general practice operates predominantly under a fee-for-service model and research participation is not routinely embedded within primary care. Patients may attend any practice and consult any GP, who can prescribe up to 12 months antidepressant medication without interim review. Mini dose formulations suitable for tapering are not readily available; escitalopram is the only antidepressant available as a commercially manufactured liquid formulation and compounded preparations incur out-of-pocket costs for patients.

Patients were recruited between March 2023 and November 2024 by searching the practice electronic health record database using a remote data extraction tool to identify and create a list for each GP of their potentially eligible patients. Inclusion criteria were age 18 years or older, prescribed any of 15 antidepressants for longer than 12 months, and the indication for prescription being a mental health condition.^32^ The included antidepressants account for 94% of the Australian market.^35^ Exclusion criteria included ‘under the care of a psychiatrist’ and ‘living in residential care’.^32^ The GPs were asked to review their list of potentially eligible patients and deselect patients that did not fit the eligibility criteria or they considered unsuitable to participate. Patients were invited via phone call to participate in the study and enrolled regardless of their intention to stop antidepressants or their baseline depression or anxiety symptom scores.^32^

Randomisation was at the general practice level and occurred after completion of patient recruitment and baseline data collection which continued for four weeks within each practice. Randomisation was via a remote central web-based randomisation service in a 3:2:2 ratio for Usual care:RELEASE:RELEASE+. Practices were stratified by size (>5/<u><</u>5 full-time equivalent [FTE] GPs). Neither patients nor GPs were blinded after allocation.

### Trial intervention

The RELEASE intervention was a low-intensity intervention comprising invitation to GP antidepressant review combined with the portable document format (pdf) RELEASE resources (https://www.releasetoolkit.com.au).^32^ The intervention included no psychological support or structured follow-up. Importantly, all patients remained under the care of their GP, and all prescribing decisions were made as usual by patient and GP together. Patients from practices randomised to the intervention were sent, via both post and email, an invitation from their GP to schedule and attend GP antidepressant review combined with the RELEASE resources including a decision aid ‘*Is stopping antidepressants right for me?*’, information brochures ‘*Stopping antidepressants’* and ‘*How family and friends can help*’, and hyperbolic tapering plans for their antidepressant.^31^

The RELEASE tapering plans provide step-by-step instructions for hyperbolic dose reduction to guide stopping over many weeks or months using antidepressant mini doses that were not provided as part of the trial. The plans were designed with safety in mind and recommend 2- to 4-weekly step-downs in dose with the flexibility to speed up, slow down, pause or revert to previous dose depending on symptoms with the patient in control of their tapering speed.^31^ The plans include 10-step, 20-step, and 40-step versions for most antidepressants based on dose reduction schedules in the Maudsley Deprescribing Guidelines.^26^ A faster 10-step plan with 2-weekly dose reductions and no pauses takes 20 weeks to complete, while a 40-step plan with 4-weekly dose reductions takes around 160 weeks to complete. The plans include instructions for accessing mini doses using compounded capsules or by crushing tablets to suspend in fluid with a link to an instructional video. The plans state: “take this tapering plan to discuss with your GP”, but patients could self-initiate. Patients were sent monthly reminders to schedule and attend GP antidepressant review for up to 6-months, beyond which there was no further intervention activity. Usual costs for consultations and medication applied, including costs borne by patients for compounded mini doses.

Intervention practices received software updates to embed RELEASE antidepressant tapering plans into the practice management software and enable customised prescribing for compounded mini doses. The trial practice liaison officer provided an optional 1-hour outreach training session for GPs, and GPs were also provided access to an optional 1-hour e-learning module. Nearby community pharmacists were provided information about the trial and intervention.

In addition to the above, for practices randomised to RELEASE+ intervention, GPs received a clinical audit and feedback package for assessing practice antidepressant prescribing. A patient facing digital platform providing access to resources, messaging and e-mental health supports was planned but proved infeasible to develop within trial timelines and was not delivered.

Usual care patients received care as usual which may include no GP antidepressant review.

### Outcomes

The primary outcome was antidepressant cessation at 12 months defined as 0mg maintained for at least two weeks.^32^ Secondary outcomes were cessation at 6 months, and ≥75% dose reduction at 6 and 12 months to capture those still tapering at follow-up. Dose reduction was considered more realistic than cessation within trial timelines.^36^ Patients who switched drugs but remained on a standard dose were considered as not achieving dose reduction. Participants with data for medication but not dose were included for cessation outcome but considered as missing for ≥75% dose reduction outcome. Secondary symptom outcomes measured at 6- and 12-months included depression (PHQ-9), anxiety (GAD-7), and withdrawal symptom scores using a pre-published version of the Discriminatory Antidepressant Withdrawal Symptoms Scale (Supplementary Appendix).^32, 37^

### Adverse events

An incident reporting proforma was developed for GPs to record and report adverse events to the Data Monitoring and Safety Committee.^32^ Any patient who recorded a score of ≥15 on the GAD-7 or the PHQ-9, or above 1 (i.e. 2 or 3) on the ninth question of the PHQ-9 survey (about suicide/self-harm) at any of the three data collection timepoints was sent an automated message advising that their score was in the range where it was recommended they schedule an appointment to see their GP.

### Sample size

Based on a previous study, we anticipated 12% and 30% of patients would cease antidepressants at 12 months in the usual care and the combined RELEASE groups respectively.^13^ Assuming a design effect due to clustering of 1.57 (intracluster correlation = 0.03; mean cluster size=20), 522 target patients gave >95% power to detect a difference of 18% for the primary outcome between the usual care and combined RELEASE groups (two-sided alpha=0.05).^34^

### Analytical methods

In the primary analysis, between-group differences in medication cessation were analysed using logistic regression, with treatment group and the stratification factor (practice size) included as fixed effects, and GP practice as a random effect. Effect estimates are presented as odds ratio (OR) with 95% confidence intervals (95%CI). Secondary medication outcomes were assessed with similar logistic regression models and are reported as OR (95%CI). Secondary outcomes assessed on the interval scale at 6 and 12 months were analysed using mixed-effects linear regression with treatment group, practice size and the baseline level of the outcome included as fixed effects, and GP practice and individual patients included as random effects, the latter nested within practice. Effect estimates are presented as mean difference (MD) and 95%CI.

All outcomes were analysed according to the intention-to-treat principle at the patient level, which included all patients following practice randomisation who provided outcome data. For the main analyses missing data were not imputed and complete-case analyses were performed.

To test the sensitivity of findings to missing data multiple imputation by chained equations was undertaken. Imputation variables included in the model were age, gender, education, smoking status, alcohol use status, years using antidepressants, and baseline general health (PHQ) and anxiety (GAD) scores. For each outcome 20 sets were imputed before results were pooled using Rubin’s combination rules to account for missing data uncertainty. Health economic and implementation evaluations, including GP and patient interviews, will be reported separately. For secondary outcomes and subgroup analyses, formal adjustment of confidence intervals for multiplicity was not performed, and these findings should be considered as exploratory. Analyses were performed with the use of Stata statistical software, version 17.0 (StataCorp).

### Patient and public involvement

The knowledge, experiences and priorities of people with lived experience of long-term antidepressant use and withdrawal symptoms were fundamental to this research. The RELEASE intervention resources were co-developed with people with lived experience of long-term antidepressant use and withdrawal symptoms, including by means of a ‘think aloud’ study.^30^ The ongoing RELEASE Lived Experience Advisory Group (LEAG) met several times each year during the conduct of the trial, sanctioned the RELEASE intervention and recommended developing additional resources including brief explanatory videos.^31^ The RELEASE research has also been informed by the opinions of investigators with lived experience, GPs,^18, 33^ and the wider public. During the course of the trial the lead author received hundreds of email requests from both consumers and practitioners in and outside Australia for access to the RELEASE hyperbolic tapering plans. In response, despite the potential for contamination of practices and GPs in the usual care arm, we made the RELEASE resources freely available online in August 2025 towards the end of the trial (www.releasetoolkit.com.au).^31^ We monitored access to the website while the trial was ongoing to check for downloads to usual care practices and GPs and there were none. Unsolicited feedback from users assisted in modifying and updating the resources.

## Results

### Practice and patient recruitment

Of 82 practices invited, 26 (32%) research naïve practices involving 195 GPs (124 full-time equivalent GPs) agreed to participate (see Figure 1). The most common reasons given for declining to participate was “GPs have no time”, followed by confidentiality concerns. Across the 26 practices, 12,069 patients were prescribed an antidepressant within 6-months, of whom 6,081 (50%) were long-term users and potentially eligible. The remote data extraction tool excluded 2,418 (40%) patients because they did not fit eligibility criteria or ‘reason for prescription’ was either absent or not a mental health diagnosis, and 26 were excluded as they had opted out of receiving SMS from the practice. A further 2,128 (35%) were excluded at the GP level, 1044 because the GP did not review their list and a further 1,088 were deselected by the GP. Reasons for deselection included ‘did not fit eligibility criteria’ (N=209) for example, under care of psychiatrist or other doctor (N=103), left the practice (N=21), or living in residential care (N=20); and “patient not suitable for invitation, for example recent personal crisis” (N=265); while for 614 no reason was given. Practices sent an SMS to the remaining 1,507 eligible patients advising of the opportunity to opt out of a pending phone call invitation into the study, which was accepted by 35 (2%). The remaining 1,472 were telephoned. Of the 1,079 (73%) who answered the phone call, 388 (36%) declined, 208 (19%) expressed interest but did not proceed, and 485 (45%) enrolled. There were two post-randomisation exclusions as baseline data revealed they were ineligible (i.e. not taking an eligible antidepressant), leaving 483.

**Figure 1.**
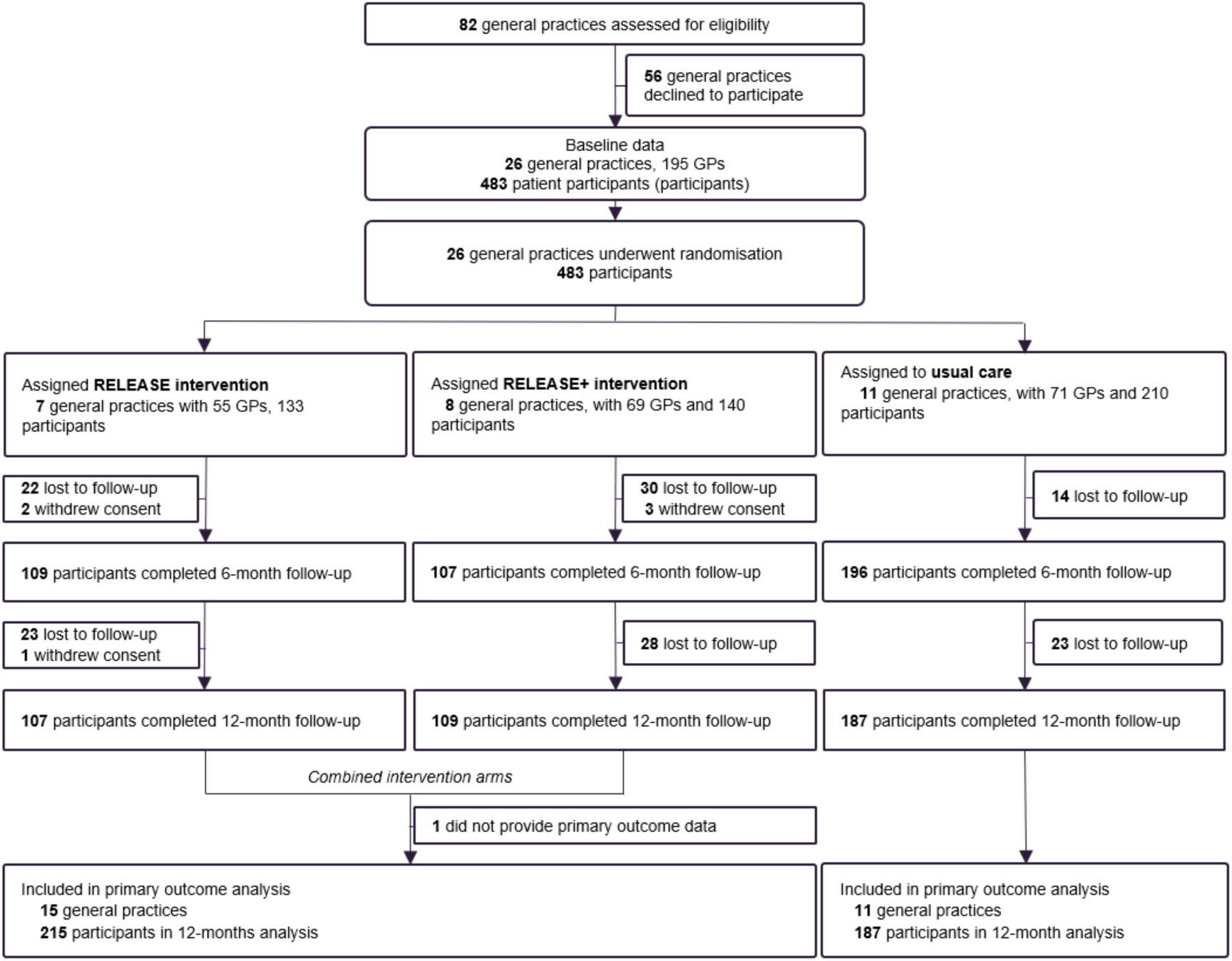
Participant flow diagram.

Eleven practices with 210 patients were randomised to usual care and 15 practices with 273 patients to the intervention (7 practices with 133 patients to RELEASE; 8 practices with 140 patients to RELEASE+).

### Baseline patient characteristics

Demographic and clinical characteristics of patients are shown in Table 1. Most were women (352; 73%), with a mean (±SD) age of 50±15 years, and average duration since first prescribed an antidepressant of 14.1±10.1 years. The most common antidepressants were escitalopram (112; 23%), sertraline (103; 21%) and venlafaxine (48; 10%). At baseline, 239 (50.7%) had no clinical indication for continued use (<10 score on both PHQ-9 and GAD-7), 55% had previously tried and failed to stop antidepressants, and only 6.2% disagreed with the statement that “my antidepressant corrects a chemical imbalance in my brain” (Table 1). Follow-up data were available for 79% of intervention and 89% of usual-care participants, with baseline characteristics of those lost to follow-up similar to those with primary outcome data (Supplementary Appendix).

**Table 1.**
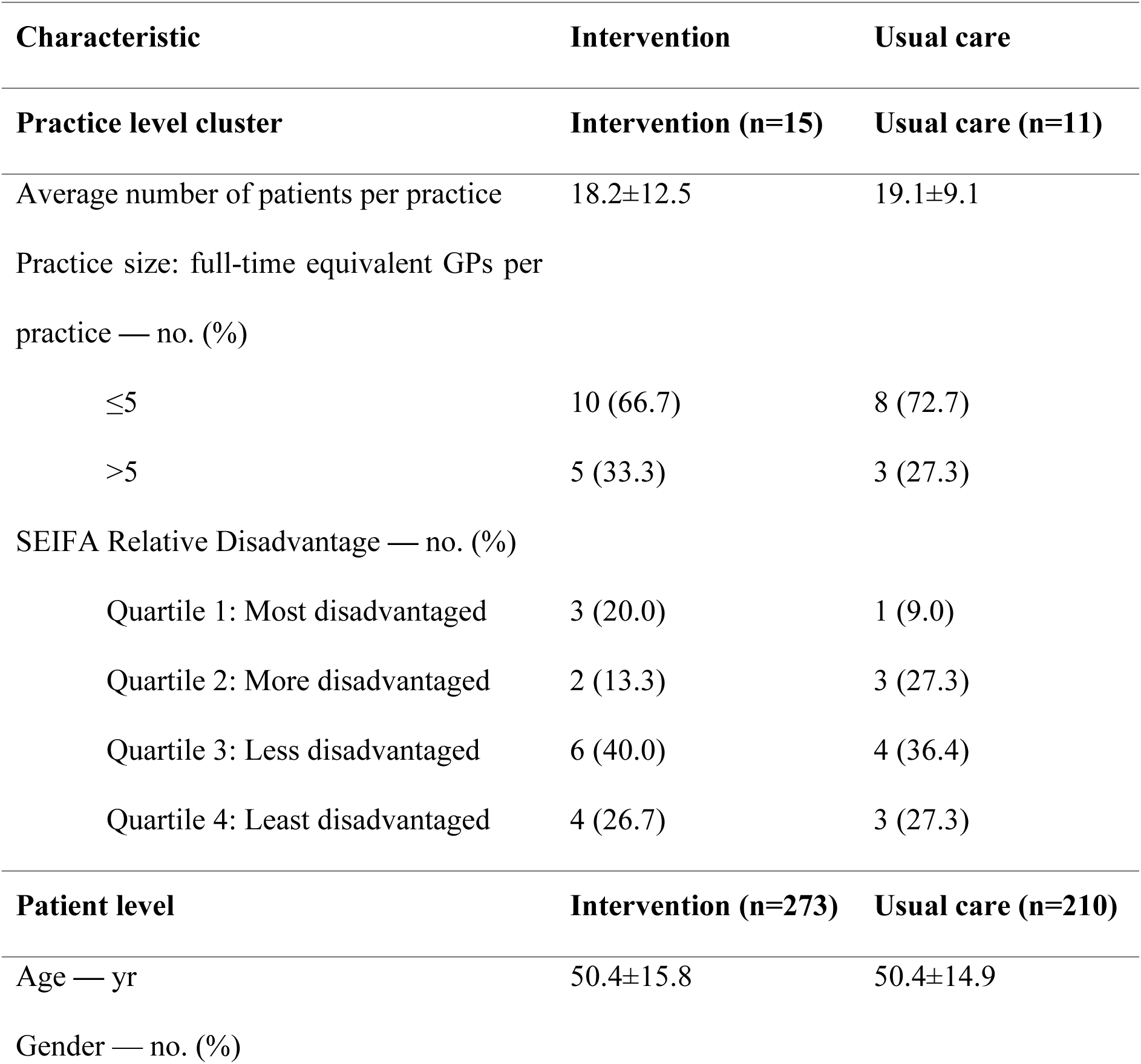
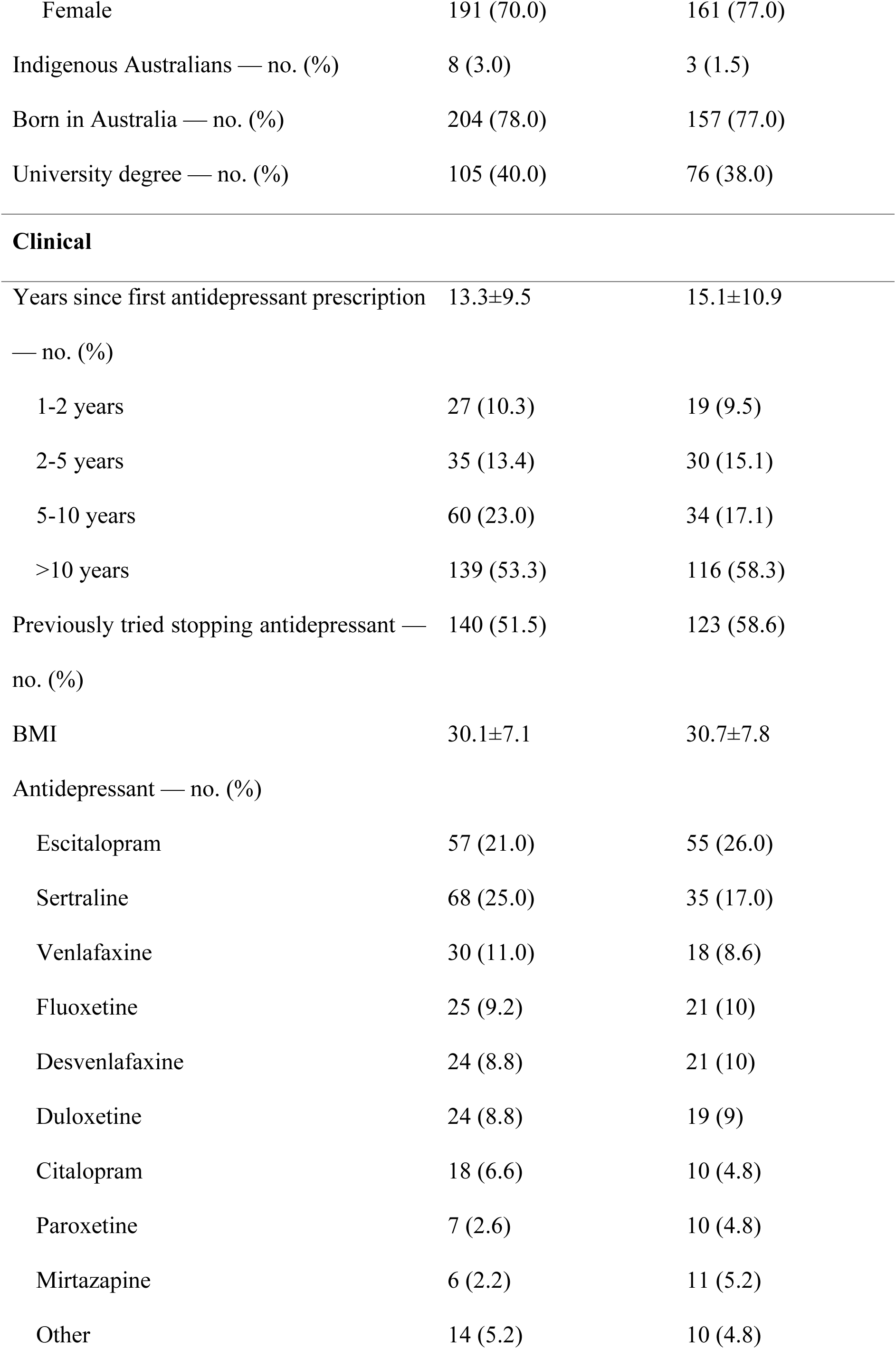
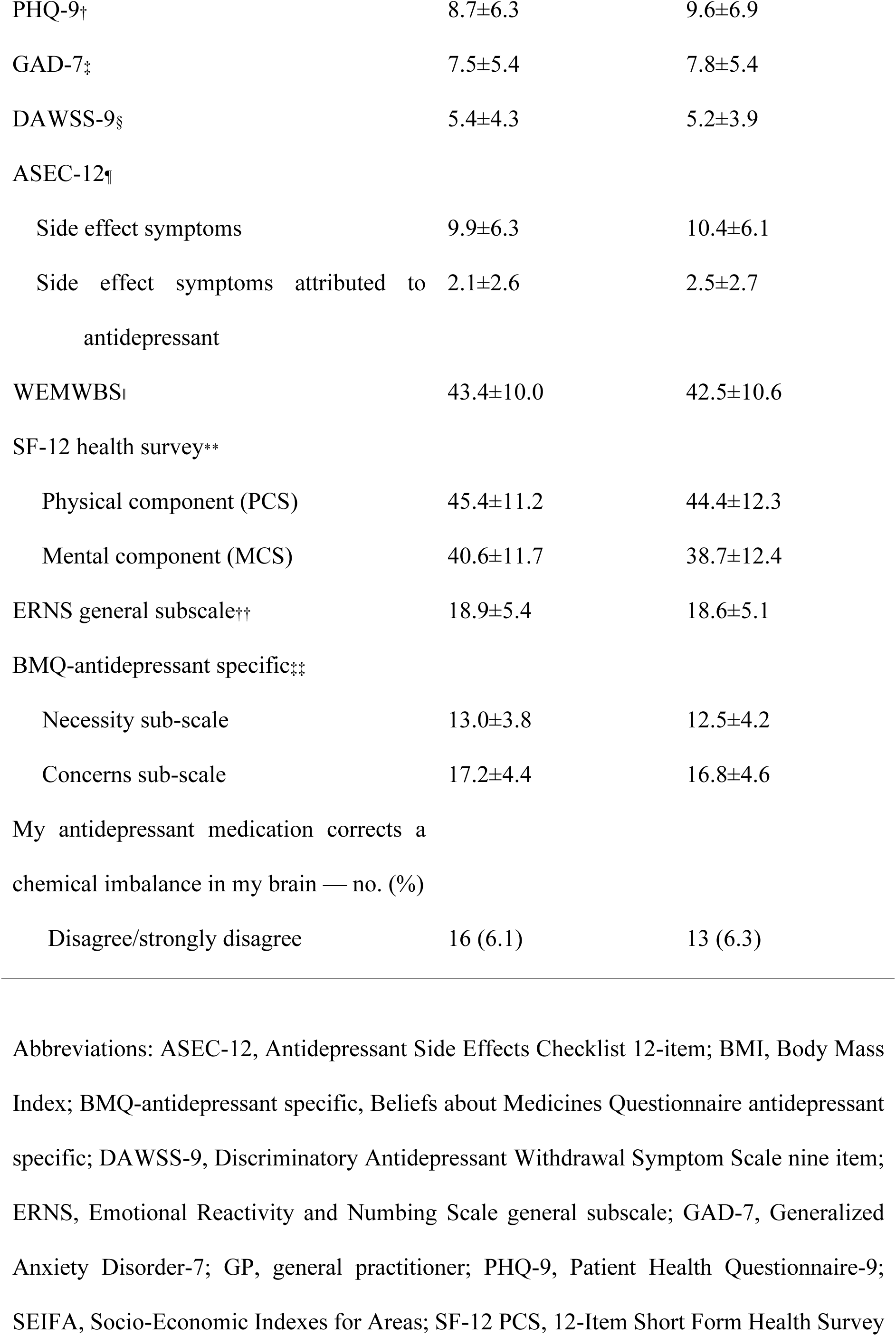
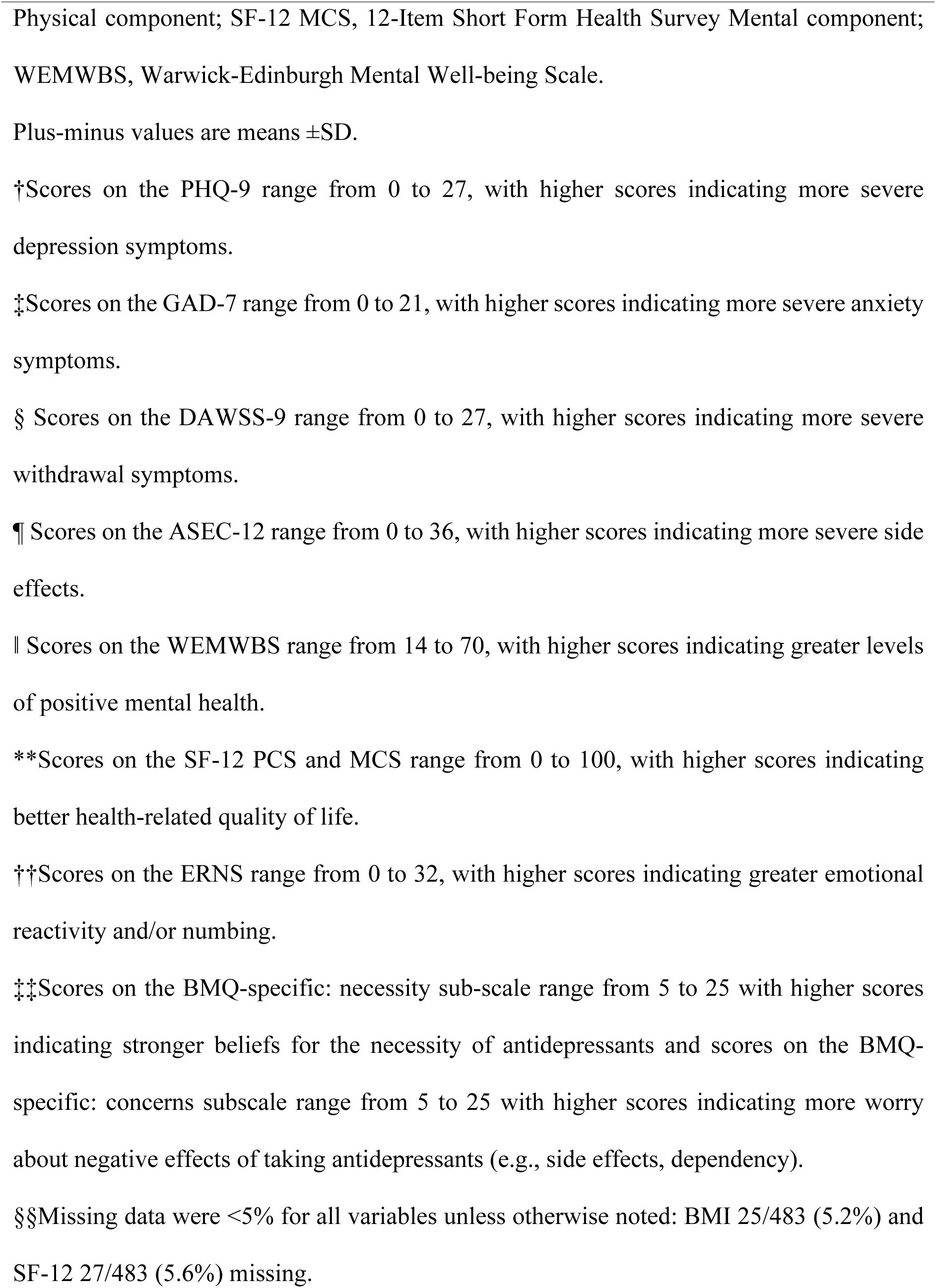
Characteristics of practices and patients at baseline.

### Primary outcome

Cessation at 12 months was observed in 32 of 215 (14.9%) intervention and 16 of 187 (8.6%) usual care patients (OR = 1.95 [95%CI, 1.00 to 3.81]; p=0.050) (Table 2). This effect is equivalent to an absolute risk difference = 6.9 percentage points [95% CI, 0.0 to 13.7] and number-needed-to-treat-to-benefit = 14 (95% CI 7 to infinity). Of those who had stopped at 12 months, more than 90% had stopped for more than 8 weeks.

**Table 2.**
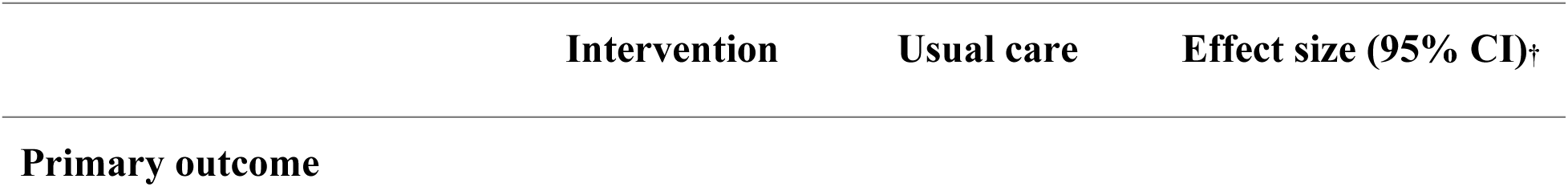
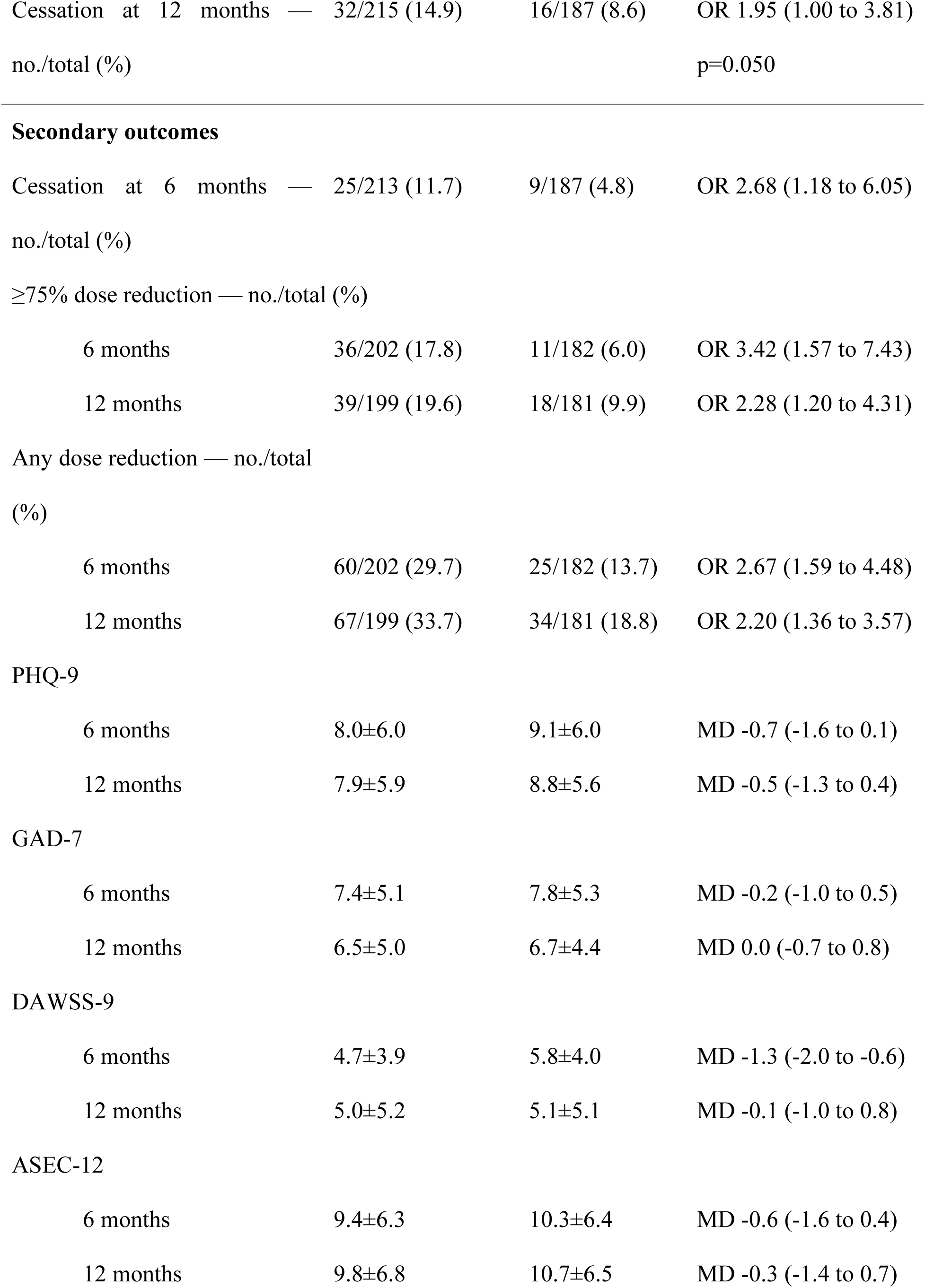
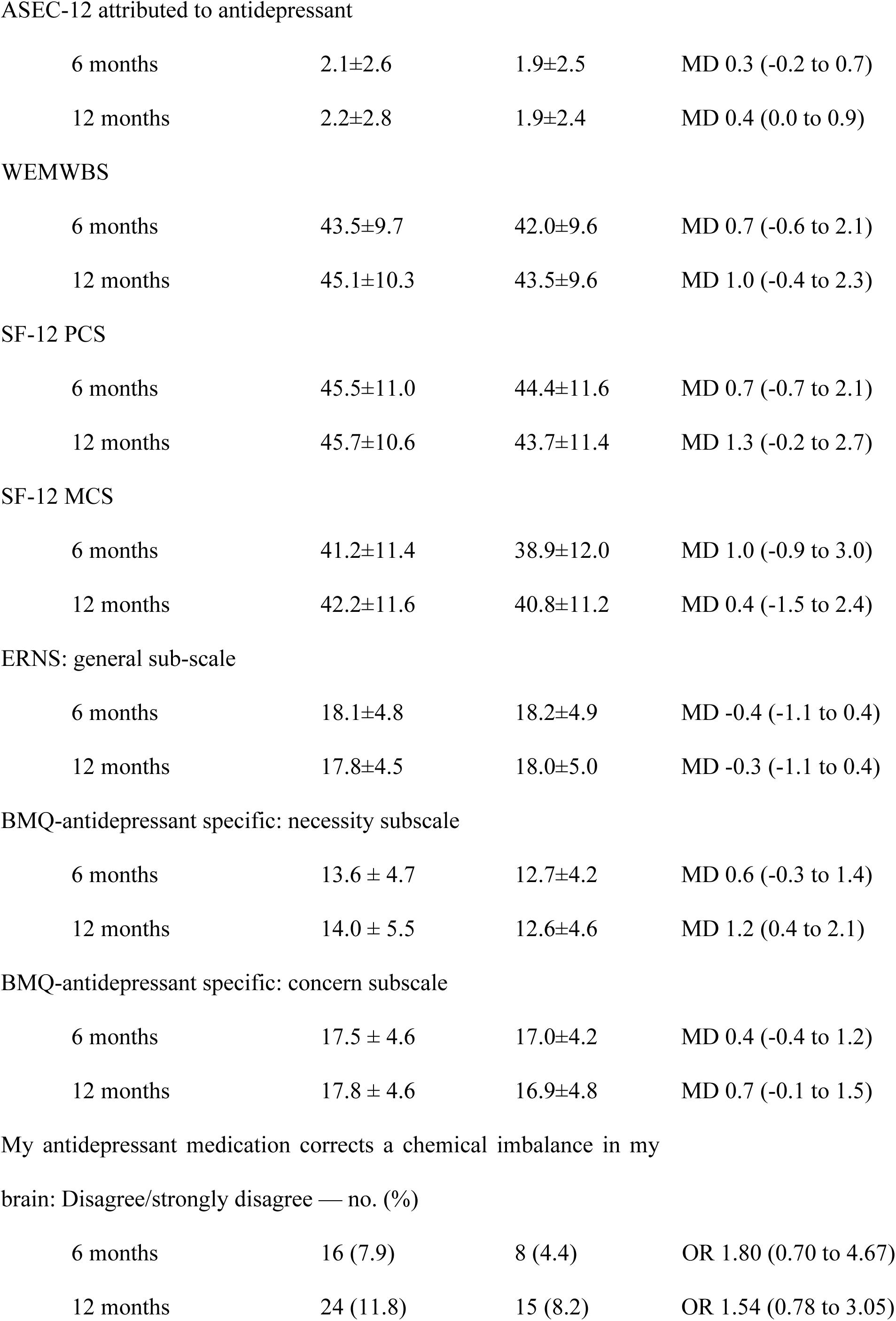
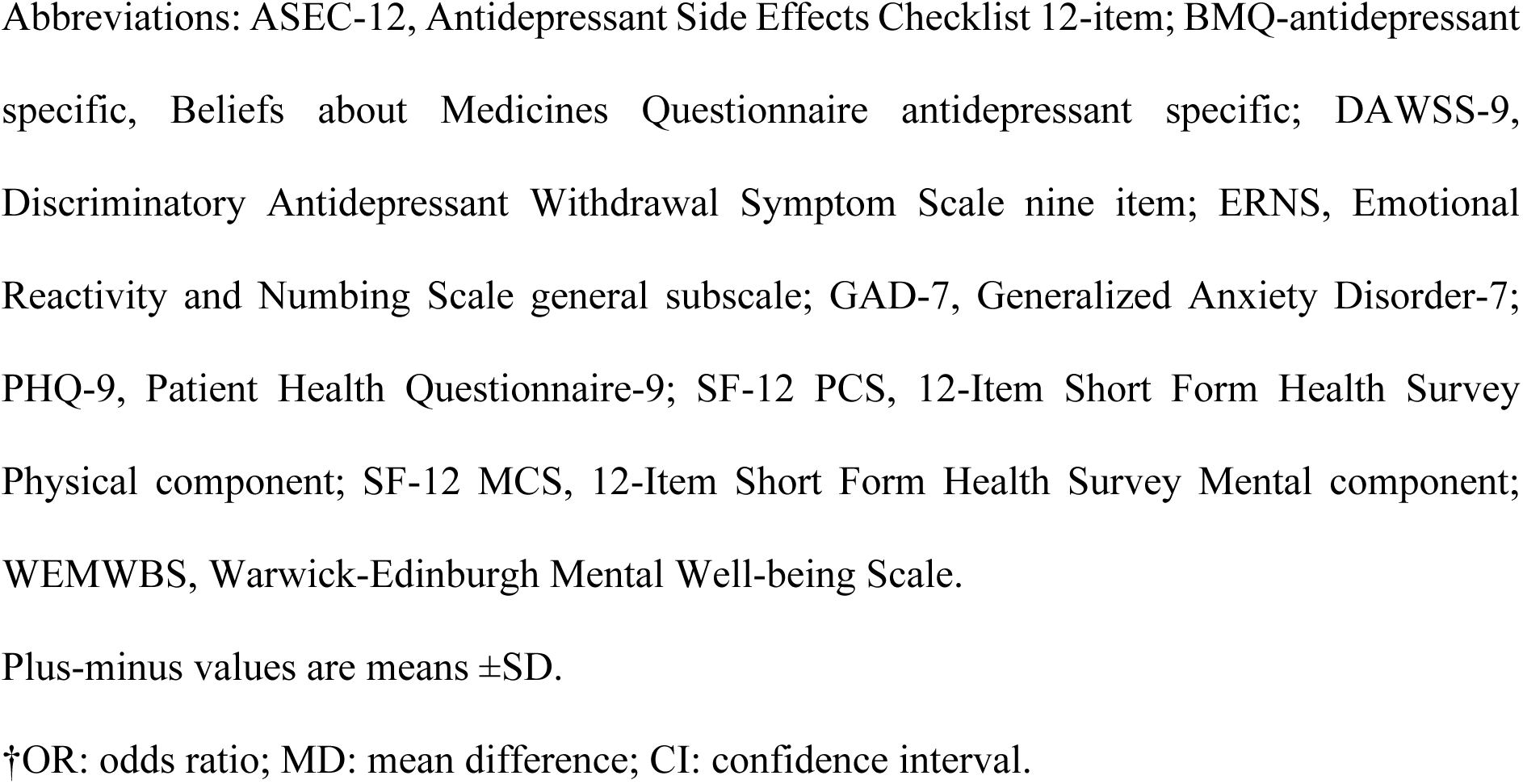
Primary and secondary outcomes.

|  | Intervention | Usual care | Effect size (95% CI)† |
| --- | --- | --- | --- |
| **Primary outcome** |  |  |  |
| Cessation at 12 months — no./total (%) | 32/215 (14.9) | 16/187 (8.6) | OR 1.95 (1.00 to 3.81)<br>p=0.050 |
| SF-12 PCS |  |  |  |
| 6 months | 45.5±11.0 | 44.4±11.6 | MD 0.7 (-0.7 to 2.1) |
| 12 months | 45.7±10.6 | 43.7±11.4 | MD 1.3 (-0.2 to 2.7) |
| My antidepressant medication corrects a chemical imbalance in my brain: Disagree/strongly disagree — no. (%) |  |  |  |
| 6 months | 16 (7.9) | 8 (4.4) | OR 1.80 (0.70 to 4.67) |
| 12 months | 24 (11.8) | 15 (8.2) | OR 1.54 (0.78 to 3.05) |

Abbreviations: ASEC-12, Antidepressant Side Effects Checklist 12-item; BMQ-antidepressant specific, Beliefs about Medicines Questionnaire antidepressant specific; DAWSS-9, Discriminatory Antidepressant Withdrawal Symptom Scale nine item; ERNS, Emotional Reactivity and Numbing Scale general subscale; GAD-7, Generalized Anxiety Disorder-7; PHQ-9, Patient Health Questionnaire-9; SF-12 PCS, 12-Item Short Form Health Survey Physical component; SF-12 MCS, 12-Item Short Form Health Survey Mental component; WEMWBS, Warwick-Edinburgh Mental Well-being Scale.
Plus-minus values are means ± SD.
†OR: odds ratio; MD: mean difference; CI: confidence interval.

Subsequent to this analysis, the RELEASE+ and RELEASE arms were compared, with cessation at 12 months observed in 15 of 108 (13.9%) and 17 of 107 (15.9%) patients respectively (OR = 0.89 [95%CI 0.37 to 2.15]), validating our design approach to combine RELEASE and RELEASE+ for the main analyses (Supplementary Appendix). Findings were similar after multiple imputation for missing data (OR = 1.82 [95%CI, 0.96 to 3.45]; Supplementary Appendix).

### Secondary outcomes

The intervention group was more likely than the usual care group to report cessation at 6 months (OR = 2.68 [95%CI 1.18 to 6.05]), dose reduction of at least 75% at both 6 (OR 3.42 [1.57 to 7.43]) and 12 months (OR 2.28 [1.20 to 4.31]) (see Table 2), and any dose reduction at 12 months (OR = 2.20 [1.36 to 3.57]) (See Figure 2). Symptom scores, including depression, anxiety and withdrawal symptom scores, remained largely similar between groups, although withdrawal symptoms scores were slightly lower in the intervention group at 6 months (Table 2).

**Figure 2.**
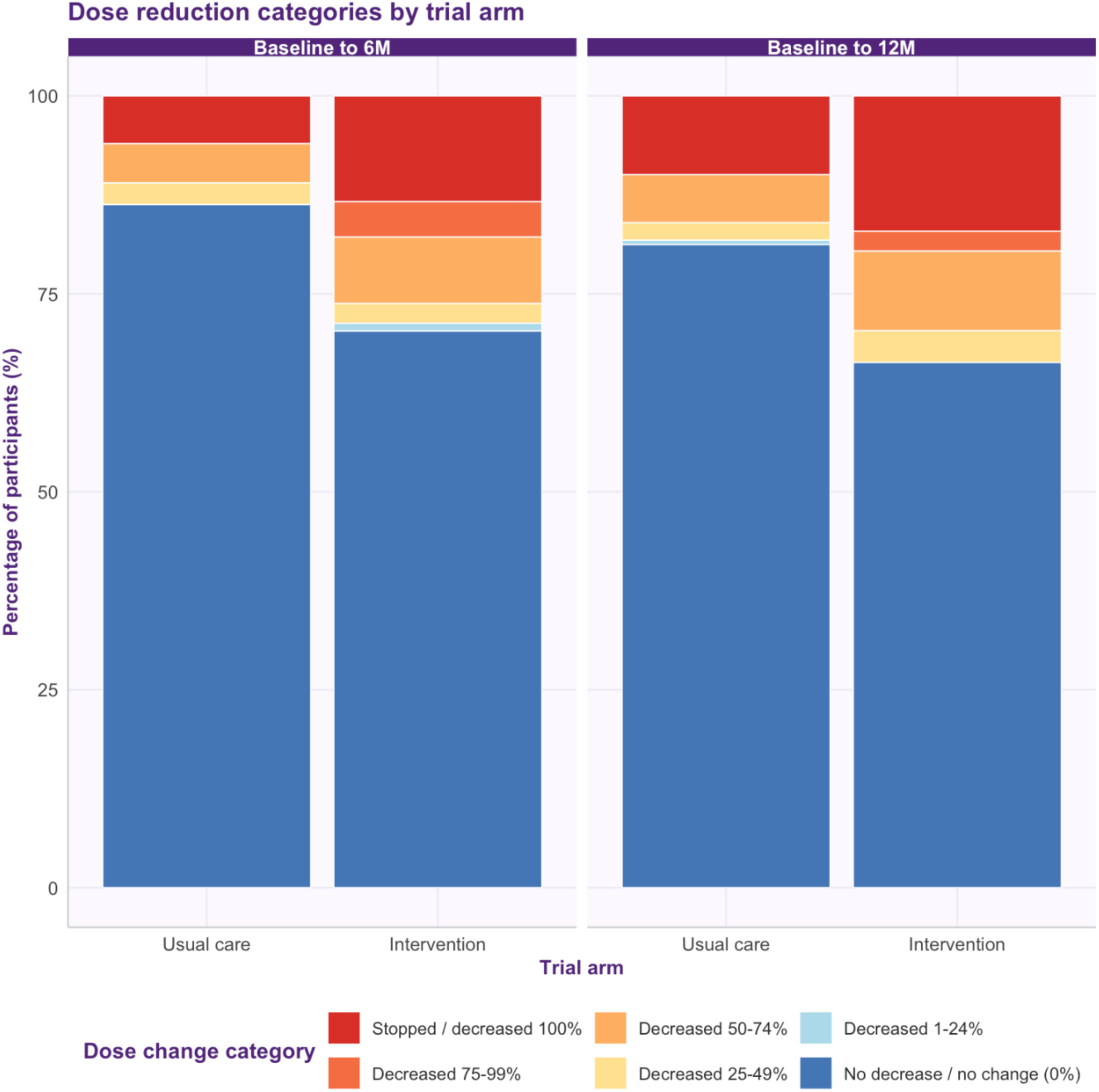
Dose reduction at 6 and 12 months comparing usual care and intervention groups.

### Intervention engagement

All 15 intervention practices were provided the outreach education and training which was attended by 64 of 124 GPs (51%), and 31 GPs (25%) accessed the e-learning module. All eight RELEASE+ practices were provided the clinical audit package, which was attended by 24 of 69 GPs (35%). By 12 months, 176 (75.2%) intervention patients had discussed stopping antidepressants with their GP versus 81 (40.3%) in usual care, and 134 (57%) had discussed a RELEASE tapering plan.

### Adverse events

No study-related adverse events were reported in participants allocated to receive the intervention. One participant died for reasons unrelated to the study.

## Discussion

In this pragmatic, cluster randomised controlled trial involving 26 general practices, people taking antidepressants long-term who received an invitation to GP antidepressant review combined with patient-centred resources to inform shared decision-making and guide hyperbolic tapering were more likely to stop or reduce antidepressant dose over 12 months compared with people receiving usual care. These changes were achieved without causing adverse effects or relapse as measured by validated depression and anxiety symptom scores.

Our effect size of around 7% absolute increase in antidepressant cessation was modest but represents a clinically meaningful effect in a challenging population with mean duration of antidepressant use of 14.1 years, in whom a low cessation rate is to be expected.^24, 37^ Based on our results, only 17 people would need to receive the low-intensity RELEASE intervention for one additional person to successfully stop antidepressants. Given the high prevalence of long-term antidepressant use, even modest effects are meaningful; for every 1000 long-term antidepressant users reached, approximately 59 additional people would stop who would not otherwise have done so. Even so, the low cessation rate in both intervention and usual care groups remains a concern given that half the cohort had no apparent indication for continued antidepressant use, a figure consistent with evidence suggesting that 30–60% antidepressant users in primary care do not meet diagnostic criteria.^38^ While modest, our results compare favourably with other trials involving patients irrespective of their intention to stop,^12^ and with discontinuation trials that found higher relapse rates although arguably mis-classified withdrawal symptoms as relapse.^22, 23^

Most patients who stopped antidepressants did so in the first 6 months suggesting an attenuation of intervention effect over time, which may be expected given there was no intervention activity in the second 6 months, and low uptake of tapering schedules that can be completed within 12 months. The proportion that had discussed stopping antidepressants with their GP (75% intervention vs. 40% usual care) and a RELEASE tapering plan (57% intervention) by 12 months stands in contrast to the proportion that achieved cessation, attesting to the challenge of stopping within 12 months and suggesting that the RELEASE intervention was more successful in prompting discussions about stopping than instituting hyperbolic tapering for successfully stopping. The RELEASE tapering plans encourage a flexible approach and can take longer than 12 months to complete. Speculation that there was low uptake of hyperbolic tapering to completion is supported by data showing that at both 6- and 12-months the proportion that achieved ≥75% dose reduction was only slightly higher than the proportion that achieved cessation, and that more achieved ≥75% dose reduction at 6-months than cessation at 12-months. It appears that patients did not progress through the final 25% of tapering schedules which involve small dose reductions through very low doses to cessation. This is consistent with clinical experience and pharmacological evidence that the second half of tapering schedules are the most difficult to achieve due to withdrawal symptoms and difficulty accessing the requisite mini doses. Our qualitative work identified poor access to mini doses (compounded capsules are expensive in Australia and crushing and dissolving tablets every day is daunting) and the belief held by both GPs and patients that tapering through low doses is unnecessary as major barriers to hyperbolic tapering.^39^

Symptom scores remained largely similar between groups across 12-months follow-up, possibly because most patients in both groups made no change in medication. At 6-months, withdrawal symptom scores were slightly lower in the intervention group compared to usual care, but likely not clinically meaningful.

### Strengths and limitations

Strengths of this study include its pragmatic design addressing an important clinical problem in primary care, the novel low-intensity intervention, and our recruitment strategy. We drew from a broad primary care population including research naïve practices and those in disadvantaged areas,^40^ and in contrast to prior trials we included patients irrespective of their intention to stop, baseline symptom scores, or antidepressant type and withdrawal symptom risk.^12, 14, 22^ Our approach enabled one in three eligible primary care patients to receive the intervention, attesting to its generalisability. Of eligible patients invited to participate via phone call, almost half enrolled - a proportion higher than the low participation rates typically observed in primary care mental health trials.^12, 14, 41^ Involving patients regardless of their intention to stop made the trial more representative of everyday general practice where many patients are unaware that stopping may be an option,^30^ but may have contributed to our low absolute cessation rate.

The focus of the low-intensity RELEASE intervention was on recognition and management of withdrawal symptoms, driven by people with lived experience of long-term antidepressant use and withdrawal symptoms. Our priority was to develop a safe, scalable approach suitable for real-world primary care where long-term antidepressant prescribing is common and rising. We defined cessation as maintained for at least two weeks, rather than many months, because the RELEASE tapering plans were designed to taper slowly over many weeks or months and through very low doses to prevent temporary cessation. While patients may restart antidepressants at any time, more than 90% of those who had stopped at 12 months had stopped for longer than 8 weeks. Our 12-month follow-up was not long enough to capture cessation in those following longer tapering schedules, but feasible during trial timelines and at least six months since any intervention activity. We included dose reduction as a secondary outcome to capture patients still tapering at follow-up. Data collection at only 6- and 12-months was insufficiently frequent to detect symptoms associated with 2- to 4-weekly dose reductions. Longer-term and more frequent follow-up are needed.

Important limitations warrant consideration. The primary outcome is clinically meaningful but borderline statistically significant. Although our sample size was well-powered based on initial calculations for a two-group comparison, the observed effect size was smaller than expected. Note that the sample is robustly sized relative to existing literature and one of the largest studies investigating strategies to address rising long-term antidepressant use in primary care.^12–14^ Differential attrition between study arms may also have influenced results. We do not have data on reasons for dropout, although baseline characteristics of those lost to follow-up were similar, and findings of our sensitivity analyses support our main findings.

Selection and volunteer biases may limit generalisability of findings. Only one third of invited practices participated. Practices with GPs more motivated to address antidepressant overuse may have been more likely to participate. Only one quarter of long-term antidepressant users were identified as eligible. This is probably an underestimate relating to both the nature of the electronic health record often lacking ‘reason for prescription’, and to our recruitment strategy providing GPs with the opportunity to review and exclude patients and our further excluding patients on lists not reviewed by GPs. While patients may have been more likely to participate if they were motivated to stop, our low absolute cessation rates compared to trials targeting patients motivated to stop suggest this was not the case.^13, 14^

### Implications for clinical practice and future research

The low-intensity RELEASE intervention is consistent with good practice and highly scalable given it rests on inviting patients to GP review combined with providing pdf resources and includes no structured follow-up or psychological therapy. The multifaceted RELEASE intervention targets both GPs and patients. We do not know which components of RELEASE contributed most to the observed effect. However, both practice participation and GP engagement with education and training opportunities were low, raising questions about feasibility of the intervention in every practice. By 12 months, 75% intervention patients had discussed stopping antidepressants with their GP and 57% had discussed a RELEASE tapering plan leading us to speculate that empowering patients with patient-centred resources, including invitation to GP antidepressant review and a clear tapering strategy, may be a key driver of effect.^39^ Patient interest in cessation could be leveraged to drive change in GP prescribing behaviour in any primary care system where invitation to review and provision of information and pdf resources are feasible.

The question of effectiveness of hyperbolic tapering for stopping long-term antidepressants remains unanswered. The low cessation rate raises the question whether future efforts should focus more strongly on challenging beliefs and increasing patient motivation and readiness to stop, in addition to providing guidance on tapering. Future research could explore whether repeated mailouts of information and resources or more support to guide tapering improve cessation rates. Scalability and broader implementation will be influenced by access to mini dose formulations, which are differentially available in different healthcare systems. Notably, antidepressant mini doses are more widely available in both the US and the UK where most antidepressants are available in liquid formulation making hyperbolic tapering more feasible.

### Conclusions

The RELEASE trial demonstrates that inviting patients to GP antidepressant review combined with patient-centred resources to inform and guide hyperbolic tapering, can support cessation or dose reduction without increasing adverse effects including relapse. Absolute cessation rates were modest but meaningful given the prevalence of potentially unnecessary long-term antidepressant use in primary care. Further research is needed to address barriers to discontinuation. These findings highlight the potential for scalable, patient-centred deprescribing approaches to mitigate the growing burden of potentially unnecessary long-term antidepressant use in primary care.

## Data Availability

All data produced in the present study are available upon reasonable request to the authors

## Acknowledgements

We thank all participating patients, general practices and general practitioners for their involvement in the RELEASE trial. We also thank the members of the RELEASE Lived Experience Advisory Group and the RELEASE Steering Committee.

## Competing interests

Katharine Wallis declares receiving reimbursement of travel expenses to attend the European College of Neuropsychopharmacology congress 2025.

Joanna Moncrieff declares grants from the National Institute for Health Research, Royalties for books about psychiatric drugs, and being co-chairperson of the Critical Psychiatry Network.

Mark Horowitz declares Royalties from the Maudsley Deprescribing Guidelines: Antidepressants, Benzodiazepines, Gabapentinoids and Z-drugs, consulting fees and stock from Outro Health a digital clinic in the US that helps patients to safely stop no longer needed antidepressants, Honoraria from universities and NHS Trusts for talks on deprescribing, and expenses for attending academic conferences.

## Funding

This research was funded by the Australian Commonwealth Department of Health, Medical Research Future Fund (MRFAR000079) and the National Health and Medical Research Council (NHMRC2015744).

## Notes

### Clinical Trial

ANZCT registry identifier, ACTRN12622001379707p.

### Clinical Protocols

https://link.springer.com/article/10.1186/s13063-023-07646-w

https://www.medrxiv.org/content/10.1101/2024.10.08.24314879v1.full

### Author Declarations

The study received approval from The University of Queensland Human Research Ethics Committee (2022/HE001667).

