## Supplementary appendix for "Redressing long-term antidepressant use (RELEASE): Pragmatic cluster randomised controlled trial in general practice"

This appendix has been provided by the authors to give readers additional information about their work.

Table of contents

1. The Discriminatory Antidepressant Withdrawal Symptom Scale nine questions (DAWSS-9)
2. Table S1: Analysis of RELEASE+ vs RELEASE.
3. Table S2: Secondary outcomes including predictors of missingness.
4. Table S3: Primary and Secondary outcomes analysed using multiple imputation.

1. **The Discriminatory Antidepressant Withdrawal Symptom Scale nine questions (DAWSS-9)**

**Antidepressant withdrawal symptoms**

Have you experienced any of the following symptoms in the PAST 4 WEEKS? If so, please rate their severity.

| | Absent | Mild | Moderate | Severe |
|---|---|---|---|---|
| Dizziness/light-headedness | | | | |
| Electric zaps | | | | |
| Memory problems | | | | |
| Muscle cramps | | | | |
| Sensation of spinning/vertigo | | | | |
| Unsteady gait | | | | |
| Nausea | | | | |
| Diarrhoea/upset stomach | | | | |
| Increased dreams or nightmares | | | | |

**Table S1: Analysis of RELEASE+ vs RELEASE for cessation and ≥75% dose reduction.**

| | RELEASE+ | RELEASE | OR (95% CI) |
|---|---|---|---|
| **Stopped 6m** | N=105, 10 (9.5) | N=108, 17 (15.7) | 0.57 (0.24, 1.33); P=0.192 |
| **Stopped 12m** | N=108, 15 (13.9) | N=107, 17 (15.9) | 0.89 (0.37, 2.15); P=0.793 |
| **Reduced 6m** | N=99, 17 (17.2) | N = 103, 21 (20.4) | 0.83 (0.38, 1.81); 0.635 |
| **Reduced 12m** | N = 97, 23 (23.7) | N=102, 21 (20.6) | 1.20 (0.61, 2.34); P=0.603 |

**Table S2: Association between participant characteristics and 12-month drop-out**

| Characteristic | Data not available at 12m (N=81) | Data available at 12m (N=402) | Effect estimate (95%CI) |
|---|---|---|---|
| Age (years), mean(SD) | 51.1 (16.4) | 50.4 (15.0) | MD = 0.7 (-2.9, 4.4) |
| Female gender, n(%) | 54 (66.7) | 298 (74.1) | OR = 0.70 (0.42, 1.17) |
| Married/De facto, n(%) | 47 (62.7) | 250 (63.6) | OR = 1.04 (0.60, 1.78) |
| First Nations, n(%) | 2 (4.7) | 8 (2.1) | OR = 2.07 (0.34, 8.88) |
| Born in Australia, n(%) | 52 (72.2) | 306 (79.5) | OR = 0.66 (0.36, 1.25) |
| Employed full-time, n(%) | 26 (37.1) | 153 (42.5) | OR = 0.80 (0.45, 1.39) |
| University Degree, n(%) | 27 (39.1) | 136 (38.3) | OR = 1.04 (0.58, 1.81) |
| Time since first antidepressant (years), mean (SD) | 13.9 (9.3) | 14.2 (10.1) | MD = -0.4 (-3.1, 2.2) |
| Have previously tried stopping, n(%) | 35 (50.7) | 201 (56.6) | OR = 0.79 (0.47, 1.32) |
| Current smoker, n(%) | 16 (23.9) | 46 (13.1) | OR = 2.07 (1.09, 3.94) |
| BMI, mean (SD) | 29.4 (7.4) | 30.2 (7.1) | MD = -0.8 (-2.7, 1.1) |
| Patient Health Questionnaire, mean (SD) | 9.6 (7.1) | 8.9 (6.5) | MD = 0.7 (-1.0, 2.4) |
| GAD-7, mean (SD) | 8.8 (6.0) | 7.4 (5.4) | MD = 1.4 (0.0, 2.8) |
| ASEC, mean (SD) | 5.4 (3.1) | 5.8 (3.0) | MD = -0.4 (-1.1, 0.3) |
| ASEC attributable, mean (SD) | 2.2 (2.7) | 2.3 (2.6) | MD = -0.1 (-0.7, 0.5) |
| WEMWBS, mean (SD) | 33.7 (11.8) | 36.6 (9.8) | MD = -2.9 (-5.3, -0.4) |
| ERNS, mean (SD) | 17.5 (6.4) | 18.6 (5.4) | MD = -1.1 (-2.5, 0.2) |
| BMQ necessity, mean (SD) | 13.4 (5.1) | 13.9 (3.8) | MD = -0.4 (-1.4, 0.5) |
| BMQ - concerns, mean (SD) | 15.1 (5.4) | 15.2 (3.6) | MD = -0.1 (-1.0, 0.8) |

**Table S3: Association between treatment group and primary and secondary outcomes using multiple imputation**

| | Intervention (N = 273) | Usual care (N = 210) | Effect size (95% CI)† |
|---|---|---|---|
| **Primary outcome** | | | |
| Cessation at 12 months — no./total (%) | 32/215 (14.9) | 16/187 (8.6) | OR 1.95 (1.00 to 3.81) p=0.050 |
| **Secondary outcomes** | | | |
| Cessation at 6 months — no./total (%) | 25/213 (11.7) | 9/187 (4.8) | OR 2.68 (1.18 to 6.05) |
| ≥75% dose reduction — no./total (%) | | | |
| 6 months | 36/202 (17.8) | 11/182 (6.0) | OR 3.42 (1.57 to 7.43) |
| 12 months | 39/199 (19.6) | 18/181 (9.9) | OR 2.28 (1.20 to 4.31) |
| Any dose reduction — no./total (%) | | | |
| 6 months | 60/202 (29.7) | 25/182 (13.7) | OR 2.67 (1.59 to 4.48) |
| 12 months | 67/199 (33.7) | 34/181 (18.8) | OR 2.20 (1.36 to 3.57) |
| PHQ-9 | | | |
| 6 months | 8.0±6.0 | 9.1±6.0 | MD -0.7 (-1.6 to 0.1) |
| 12 months | 7.9±5.9 | 8.8±5.6 | MD -0.5 (-1.3 to 0.4) |
| GAD-7 | | | |
| 6 months | 7.4±5.1 | 7.8±5.3 | MD -0.2 (-1.0 to 0.5) |
| 12 months | 6.5±5.0 | 6.7±4.4 | MD 0.0 (-0.7 to 0.8) |
| DAWSS-9 | | | |
| 6 months | 4.7±3.9 | 5.8±4.0 | MD -1.3 (-2.0 to -0.6) |
| 12 months | 5.0±5.2 | 5.1±5.1 | MD -0.1 (-1.0 to 0.8) |
| ASEC-12 | | | |
| 6 months | 9.4±6.3 | 10.3±6.4 | MD -0.6 (-1.6 to 0.4) |
| 12 months | 9.8±6.8 | 10.7±6.5 | MD -0.3 (-1.4 to 0.7) |
| ASEC-12 attributed to antidepressant | | | |
| 6 months | 2.1±2.6 | 1.9±2.5 | MD 0.3 (-0.2 to 0.7) |
| 12 months | 2.2±2.8 | 1.9±2.4 | MD 0.4 (0.0 to 0.9) |
| WEMWBS | | | |
| 6 months | 43.5±9.7 | 42.0±9.6 | MD 0.7 (-0.6 to 2.1) |
| 12 months | 45.1±10.3 | 43.5±9.6 | MD 1.0 (-0.4 to 2.3) |
| SF-12 PCS | | | |
| 6 months | 45.5±11.3 | 44.4±11.6 | MD 0.8 (-0.7 to 2.1) |
| 12 months | 45.7±10.6 | 43.7±11.4 | MD 1.3 (-0.2 to 2.7) |
| SF-12 MCS | | | |
| 6 months | 41.2±11.4 | 38.9±12.0 | MD 1.0 (-0.9 to 3.0) |
| 12 months | 42.2±11.6 | 40.8±11.2 | MD 0.4 (-1.5 to 2.4) |
| ERNS: general sub-scale | | | |
| 6 months | 18.1±4.8 | 18.2±4.9 | MD -0.4 (-1.1 to 0.4) |
| 12 months | 17.8±4.5 | 18.0±5.0 | MD -0.3 (-1.1 to 0.4) |
| BMQ-antidepressant specific: necessity subscale | | | |
| 6 months | 13.6 ± 4.7 | 12.7±4.2 | MD 0.6 (-0.3 to 1.4) |
| 12 months | 14.0 ± 5.5 | 12.6±4.6 | MD 1.2 (0.4 to 2.1) |
| BMQ-antidepressant specific: concern subscale | | | |
| 6 months | 17.5 ± 4.6 | 17.0±4.2 | MD 0.4 (-0.4 to 1.2) |
| 12 months | 17.8 ± 4.6 | 16.9±4.8 | MD 0.7 (-0.1 to 1.5) |

| | | | |
|---|---|---|---|
| My antidepressant medication corrects a chemical imbalance in my brain: Disagree/strongly disagree — no. (%) | | | |
| 6 months | 16 (7.9) | 8 (4.4) | OR 1.80 (0.70 to 4.67) |
| 12 months | 24 (11.8) | 15 (8.2) | OR 1.54 (0.78 to 3.05) |
